# Whole-Cell Dissociated Suspension Analysis in Human Brain Neurodegenerative Disease: A Pilot Study

**DOI:** 10.1101/2021.01.08.21249455

**Authors:** Geidy E. Serrano, Jessica E. Walker, Anthony J. Intorcia, Michael J. Glass, Richard A. Arce, Ignazio S. Piras, Joshua S. Talboom, Courtney M. Nelson, Brett D. Cutler, Lucia I. Sue, Lih-Fen Lue, Matthew Huentelman, Thomas G. Beach

## Abstract

Biochemical analysis of human brain tissue is typically done by homogenizing whole pieces of brain and separately characterizing the proteins, RNA, DNA, and other macromolecules within. While this has been sufficient to identify substantial changes, there is little ability to identify small changes or alterations that may occur in subsets of cells. To effectively investigate the biochemistry of disease in the brain, with its different cell types, we must first separate the cells and study them as phenotypically defined populations or even as individuals. In this project, we developed a new method for the generation of whole-cell-dissociated-suspensions (WCDS) in fresh human brain tissue that could be shared as a resource with scientists to study single human cells or populations. Characterization of WCDS was done in paraffin-embedded sections stained with H&E, and by phenotyping with antibodies using immunohistochemistry and fluorescence-activated cell sorting (FACS). Additionally, we compared extracted RNA from WCDS with RNA from adjacent intact cortical tissue, using RT-qPCR for cell-type-specific RNA for the same markers as well as whole transcriptome sequencing. More than 11,626 gene transcripts were successfully sequenced and classified using an external database either as being mainly expressed in neurons, astrocytes, microglia, oligodendrocytes, endothelial cells, or mixed (in two or more cell types). This demonstrates that we are currently capable of producing WCDS with a full representation of different brain cell types combined with RNA quality suitable for use in biochemical analysis.

## Introduction

Biochemical analysis of human neurodegenerative brain tissue and animal models have produced much of what is known about these conditions and has led to FDA-approved therapies. The typical approach has been to homogenize whole pieces of frozen brain tissue and separately characterize the proteins, RNA, DNA, and other macromolecules within. However, many recent studies have recognized challenges in finding small biochemical changes that could occur in specific subsets of cells in the diseased brain. Furthermore, neurodegenerative disease often leads to massive losses of the disease-targeted cells; for example, the entorhinal cortex layer II stellate neurons or substantia nigra pigmented neurons. Whole-homogenate analysis of such brain regions can generate misleading results, as any biochemical constituent that is selectively localized to the depleted cells will appear to be “down-regulated.” Also a relevant loss or increase might be missed entirely if the biochemical entity is found in many cell types, diluting the ‘lost” signal from the cell of interest, especially if that cell type is uncommon or rare. To effectively investigate the biochemistry of neurodegenerative disease in the brain, with its thousands of different cell types, we must first separate the cells and study them as phenotypically-defined populations, and even as individuals.

Laser-capture microscopy (LCM) is a method that can pick individual cells from a cryostat section for characterization. However, this technology is limited by the considerable time and personnel investment as well as the limited ability to mark target cells phenotypically prior to microdissection. In recent years, methods have been developed that allow an initial creation of single-cell suspensions (Darmanis et al., 2015; Guez-Barber et al., 2012; Otero-Garcia et al., 2020) or single nuclei isolation from solid fixed, fresh or frozen tissue (Bakken et al., 2018; Blue B. Lake et al., 2016; B. B. Lake et al., 2017; Srinivasan et al., 2020) followed by the analysis of phenotypically-defined cells sorted based on cell-type identifying proteins or RNA expression.

These methods are much more time and labor-efficient than LCM and allow sorting by a much more diverse panel of markers. Studies suggest that even though comparable, nuclear mRNA is present at only 20-50% abundance compared to that present in whole cells (Bakken et al., 2018; B. B. Lake et al., 2017). Some groups have already published intriguing results from Alzheimer’s disease (AD) brain nuclei (Celis et al., 2020; Mathys et al., 2019; Srinivasan et al., 2020; Zhou et al., 2020) but to our knowledge only one study isolated whole cells from frozen human AD brains (Darmanis et al., 2015). In this study we explored a new methodology to create whole-cell-dissociated-suspensions (WCDS) from rapidly autopsied human brains, with the primary goal of sharing this new resource with researchers interested in studying cell-type-specific changes in aging and aging-related disorders. We analyzed possible changes that could be induced by cell isolation and suggest that our WCDS resource could help uncover cell-specific changes of aging.

## Material and Methods

### Whole-cell-dissociated-suspension preparation

Fresh brain samples came from subjects who were volunteers in the Arizona Study of Aging and Neurodegenerative Disorders (AZSAND) and the Brain and Body Donation Program (BBDP; www.brainandbodydonationprogram.org), a longitudinal clinicopathological study of healthy aging, cognition, and movement in the elderly since 1996 in Sun City, Arizona (Beach et al., 2015; Beach et al., 2008). All subjects signed an Institutional Review Board-approved informed consent, allowing both clinical assessments during life and several options for brain and bodily organ donation after death. Cases were selected independently of their clinical diagnosis, but favoring those with the shortest postmortem intervals. Fresh bilateral coronal sections of the frontal lobe were collected just anterior to the genu of the corpus callosum at autopsy and were stored in Hibernate A (Brain bits cat#HA) until processing (<18hrs). The grey matter was dissected and finely minced in the cold with RNAse later and weighed. Accutase (Innovative cell technologies BE cat#AT104) was used for enzymatic digestion and different incubation times were tested; 0 hrs, 2 hrs, and 4 hrs all at 4°C, followed by mechanical disruption by repetitive pipetting. Homogenates were centrifuged and Accutase was replaced by Hank’s balanced salt solution (HBSS), following cell filtration using 100 and 70 µm filters. Myelin, neuropil, and other cellular debris were removed using 30% and 70% Percoll (GE Healthcare cat#17-0891-01), while final WCDS were stored in cryoprotectant solution (90%FBA and 10% DMSO + 1U/µl RNAse inhibitor) for future characterization (Guez-Barber et al., 2012).

### Histological characterization

Single WCDS aliquots from all cases were fixed in formalin overnight. The day after, fixed WCDS were washed, pelleted, and embedded in paraffin. Serial 3 µm paraffin sections were collected for hematoxylin & eosin staining (H&E) and immunostaining for cell-specific markers. Antibodies targeting cell types included neuronal markers NeuN, MAP2, and neurofilament; astrocyte marker GFAP; and microglia markers IBA1 and LN3 (Table1). Stained sections were examined by a neuropathologist, who also estimated the percentages of different cell types by nuclear morphology in 12 WCDS (Figure 1).

**Figure 1.**
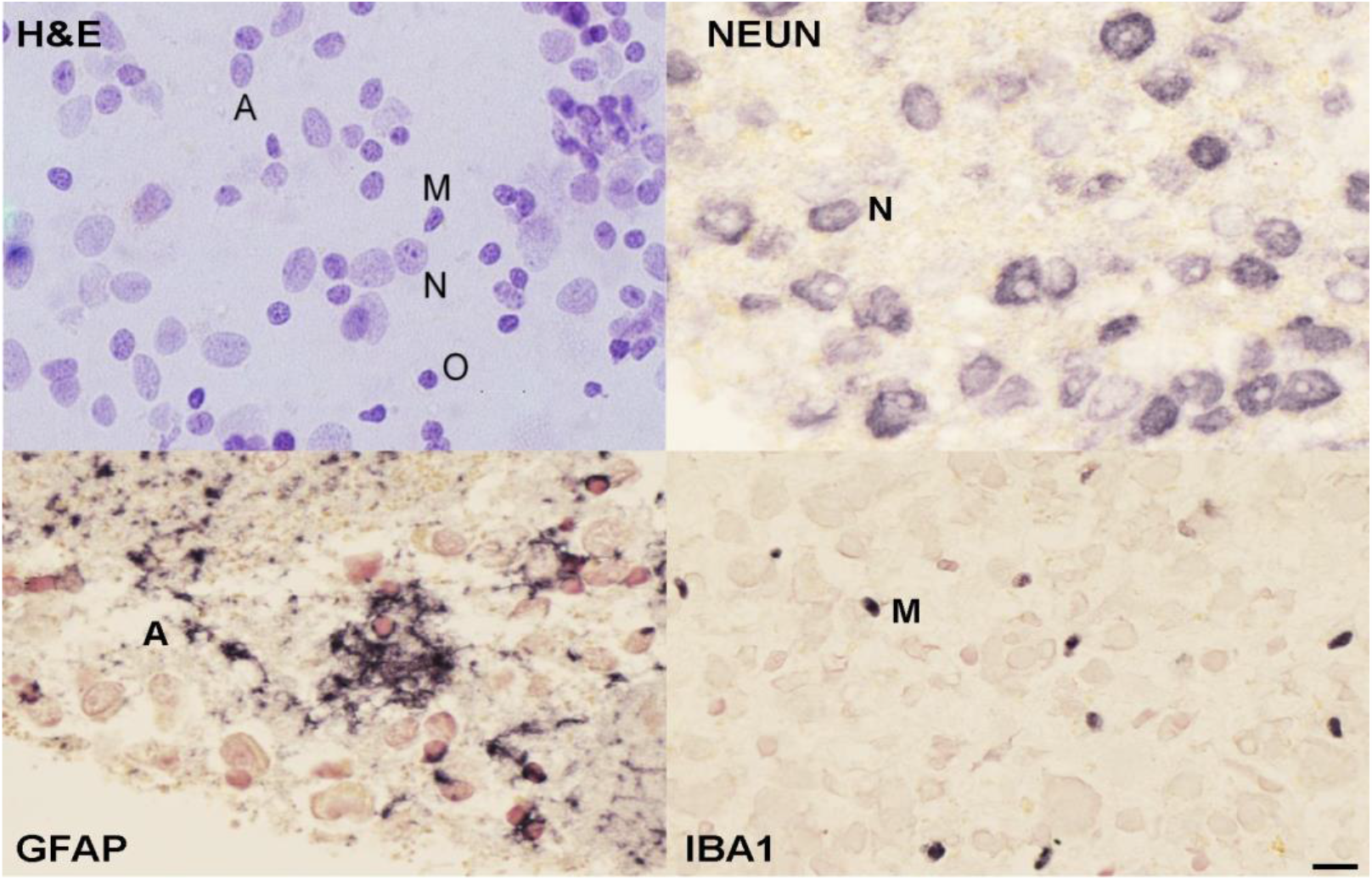
Characterization of single-cell suspensions by H & E and immunostaining with NeuN, GFAP, and IBA1). Estimation of cell numbers using nuclear morphology (H&E stain, above left) and cell-type-specific markers suggest that roughly 50% of the cells are neurons (***N=neurons***), with astrocytes (***A=astrocytes***), and microglia (***M=microglia***) each making up about 25% of the total. Oligodendrocytes (***O***) are relatively rare. *Scale bar=10µm*

### Cellular-specific marker characterization by fluorescent cell sorting

To further establish the presence of the different human brain cell types in WCDS, fluorescence-activated cell sorting (FACS; Bio-Rad S3E; with 488 and 647 nm wavelength) was used. Frozen WCDS aliquots were rapidly thawed at 37°C, followed by a 10-minute fixation in 70% methanol, 1mM EDTA, and 1U/µl of RNAse inhibitor(Alles et al., 2017; Guez-Barber et al., 2012). Suspensions were permeabilized in a 0.1 M phosphate base solution with 2% Triton X-100 for 10 minutes before primary antibody incubation overnight at 4°C. The Sorting antibodies included neuronal markers NeuN, MAP2, and neurofilament-H, GFAP marker for the activation of astrocytes, and IBA1 for microglia. Multiple antibodies tested were commercially pre-labeled with fluorophores (Table 1). Although, we also tested non-fluorescent primary antibodies, which were later incubated with fluorescent dye-labeled secondary antibodies (Molecular Probes goat anti-mouse and goat anti-rabbit 488 and 647).

**Table 1.**
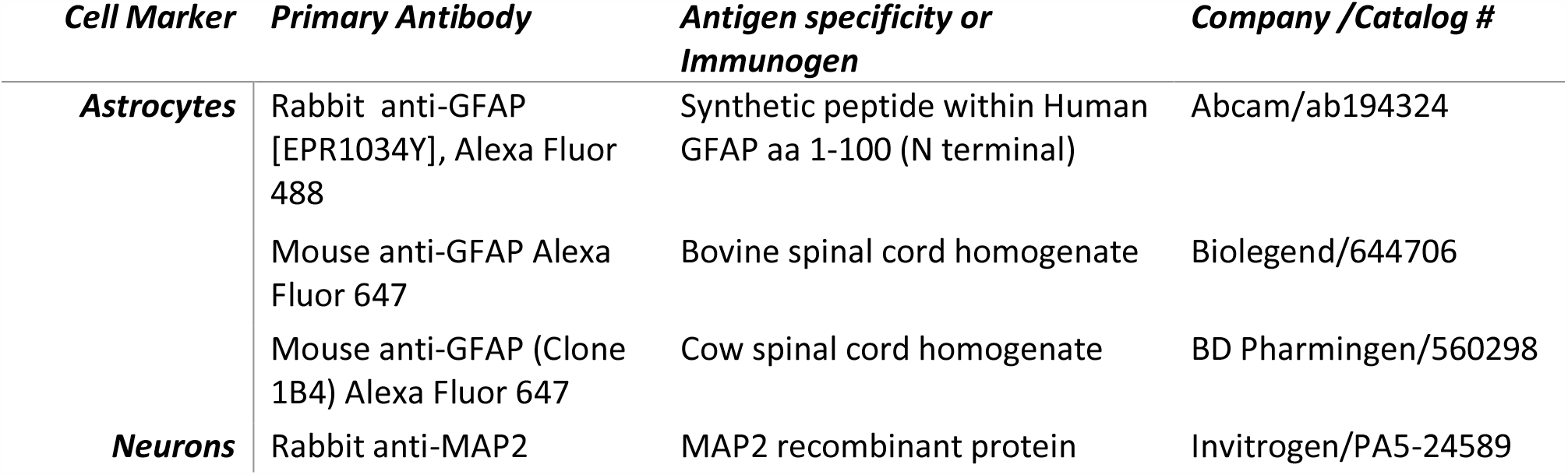
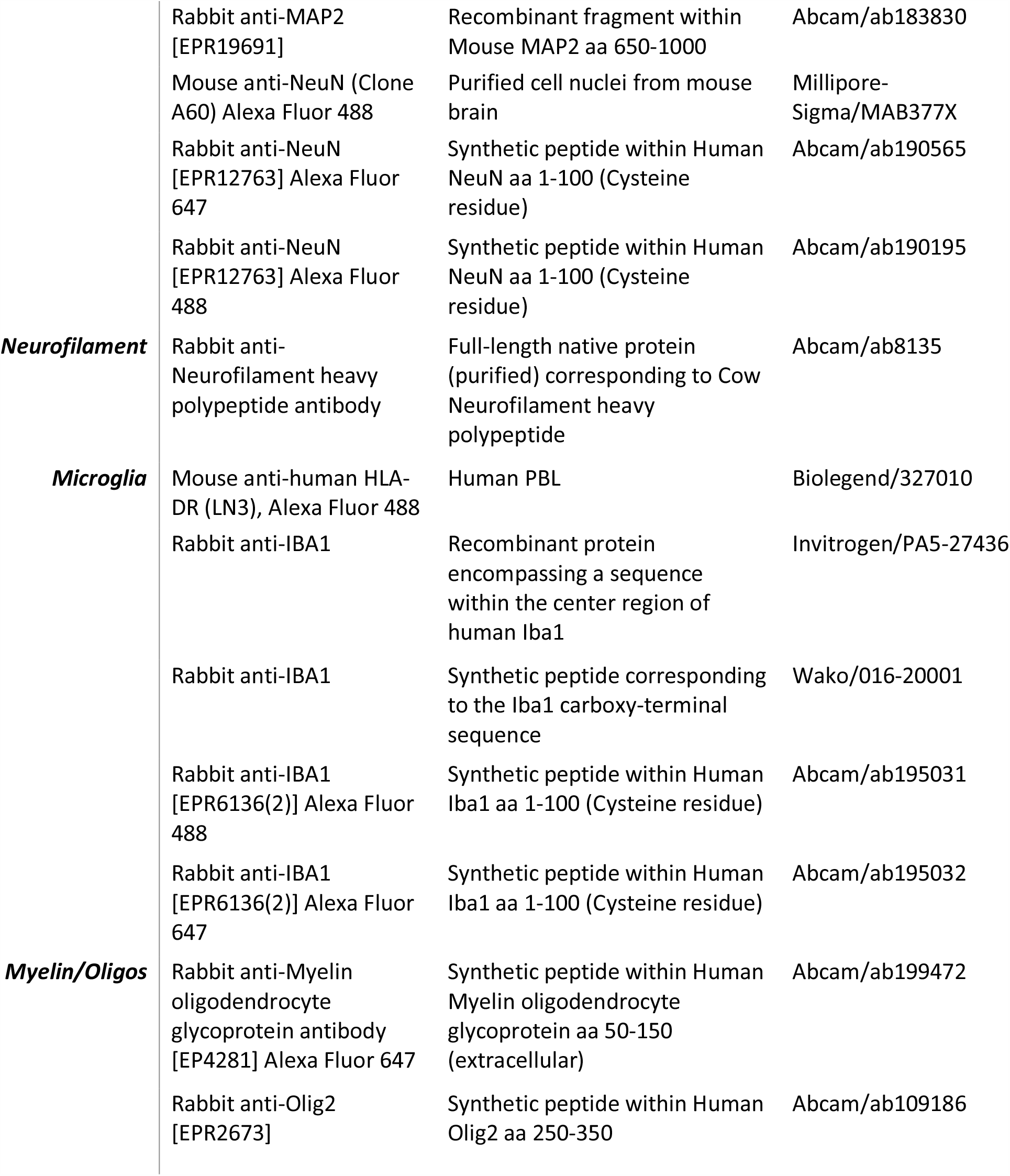
List of antibodies used to evaluate WCDS.

### RNA isolation and RNA characterization

WCDS aliquots from 30 cases were randomly selected for RNA extraction using Qiagen RNeasy Plus micro (cat#74034,) following the manufacturer’s instructions, including their suggested protocol of adding PolyA Carrier RNA. An Agilent Bioanalyzer was used to calculate the average RNA integrity number (RIN) and Thermo Fisher nanodrop to calculate the RNA yield. Following this, WCDS from 12 cases were used to further characterize the samples’ transcripts, and were then compared to those expressed in adjacent frozen whole tissue homogenates (WTH) of the same cases. RNA from WTH was extracted using Qiagen RNeasy Plus mini kits (cat# 74134). Reverse transcription for both sets of samples was done using iScript Reverse Transcription Supermix (Bio-Rad cat # 1708841). Before qRT-PCR, samples were subject to preamplification using TaqMan PreAmp Master Mix 2X (Thermo Fisher cat #4391128) and 0.2x pooled TaqMan Gene Expression Assays (Thermo Fisher cat #4351370). Probes included NeuN (Hs01370654), GFAP (Hs00909233_m1) and IBA1 (Hs00610419_g1), in addition to housekeeping probes: ACTB (Hs01060665_g1), GAPDH (Hs00266705_g1), and 18S (Hs99999901_s1). The amplification mixture was prepared using Sso Advanced Universal Probes Supermix 2X (Bio-Rad cat#1715281) and the previously mentioned TaqMan Gene Expression Assays, which include the forward and reverse primer and fluorogenic probe and preamplified cDNA. qRT-PCR was performed using Bio-Rad CFX Connect. The cycling conditions were an initial denaturation step of 95°C for 10 min, followed by 40 cycles of denaturation at 95°C for 5 sec and annealing/extension at 60°C for 1 min. A delta-delta approach was used for the qRT-PCR analysis.

### Cell sequencing

RNA extracted from the same 12 cases were used to do whole transcriptome sequencing and differential analyses, comparing WCDS versus WTH. Sequencing libraries were prepared with 100 ng of total RNA using Illumina’s Truseq RNA Sample Preparation Kit v2 (Illumina, Inc.) following the manufacturer’s protocol. The final library was sequenced by 2x 75 bp paired-end sequencing on a HiSeq 2500. After sequencing, FASTQs files were processed using STAR and HTSeq, obtaining a count table summarized at the gene level. Raw counts were filtered for genes with average counts less than five and were normalized using DESeq2 (PMID: 25516281). The differential analysis was conducted with DESeq2 using a paired design, and we selected the differentially expressed genes by FDR < 0.01 and log2FC > |1.5|, aiming to include all the genes most differentiated between the two groups (HY., 1995; Love, Huber, & Anders, 2014). We classified the genes according to their specific cell expression signatures using a single cell mRNA database from the mouse cortex. Using these data, we defined an enrichment score based on deconvolution of the known relative expression of genes in different cell types, thus assigning each gene transcript to a specific cell (neuron, astrocyte, microglia, endothelial cell, and oligodendrocytes) or a “mixed” category when the expression was not specific to any cell type (Piras et al., 2020; Zhang et al., 2016).

## Results

The resulting yield of WCDS was roughly 16 million cells per gram of fresh human gray matter. Paraffin-embedded cell pellets were stained with H&E for nuclear morphology and immunohistochemically stained with antibodies specific for neurons (neurofilament, Neu N), astrocytes (glial fibrillary acidic protein, GFAP) and microglia (Iba1) to confirm that major cell types were present. Each examined dissociated cell suspension always had a diverse population by nuclear morphology typically including approximately 40% neurons, 25% astrocytes, 21% microglia, 5% oligodendrocytes, and 4% endothelial cells (fig 1). Cell soma were intact; astrocytes occasionally had attached cell processes, while these were less common in neurons and microglia. Our cell suspension also had detached processes, but for the purpose of this study we did not attempt beyond initial centrifugation steps to further remove them from the suspensions. Cell surface and intracellular cell-specific antigen were preserved in all examined WCDS, which allowed us to also separate cell-specific populations by FACS and confirm similar percentages of neurons, astrocytes, and microglia (Fig 2).

**Figure 2.**
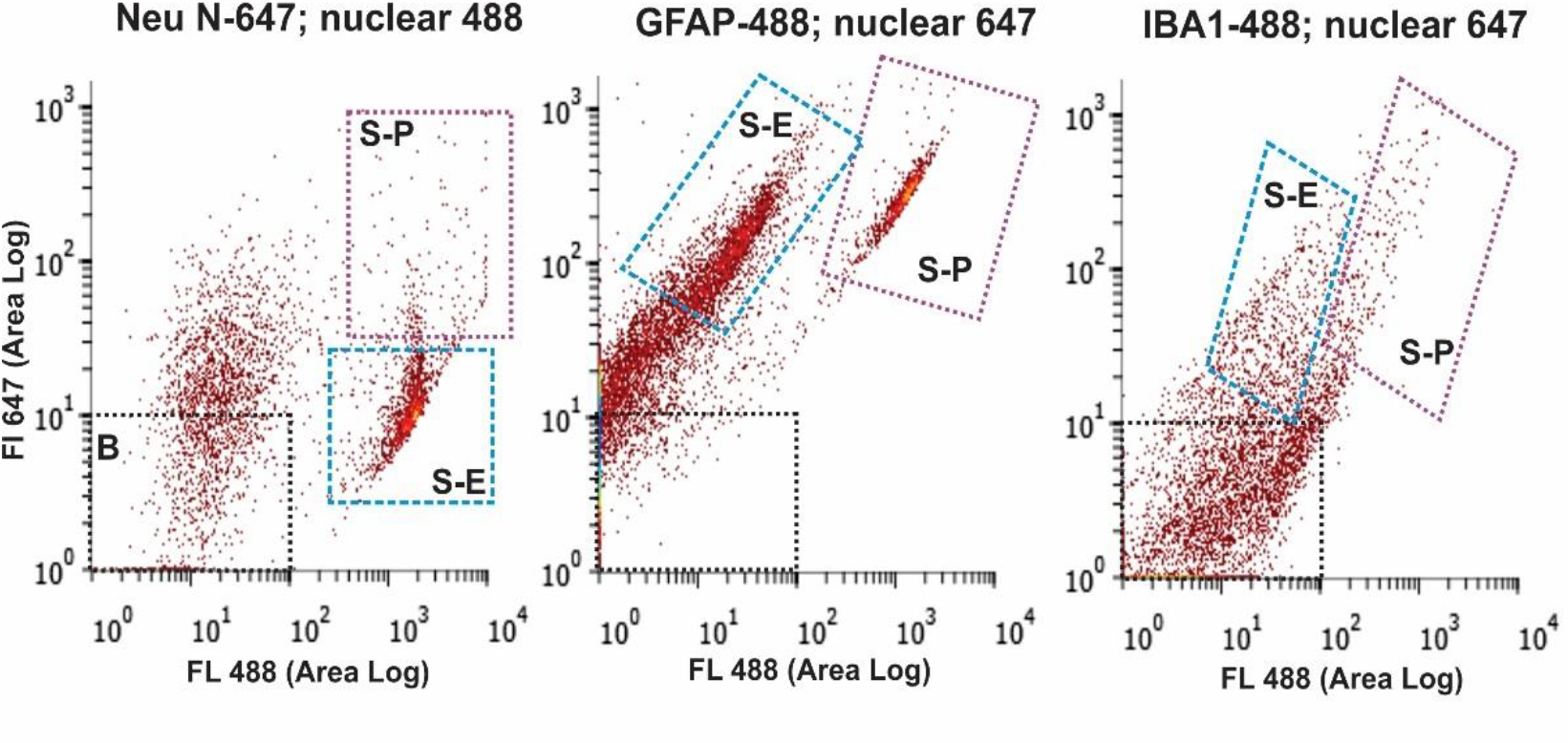
Fluorescence-activated cell sorting (FACS) of whole-cell suspensions labeled with NeuN, GFAP, and IBA1 (above). FACS allowed separation (S-P in all 3 panels above) of cells fluorescing with both a non-specific nuclear stain (488 nm, x-axis) and cell-type-specific antibodies (647 nm, Y-axis). The collection of these cells would allow further transcriptomic and proteomic analysis of specific cell populations.

Accutase enzymatic digestion was optimal at 4 hrs at 4°C, as judged by the RNA integrity and ability to create dissociated suspensions without clumps. RIN for unfixed WCDS incubated for 4 hrs in enzyme ranged from 2 to 8, with a mean RIN of 6.2 and 2.1 standard deviations, while 2 hrs incubations resulted in an average RIN of 4.8+/-3.3 and no enzyme incubation 2.7+/-0.07. The yield of RNA ranged from 4-350 ng/million cells, with a mean yield of 55 ng/million cells. Similarly, RIN from methanol-fixed WCDS that were incubated in enzyme for 4 hrs ranged from 2 to 10, with a mean RIN of 5 +/-3.6 and a yield of 60 ng/million cells +/-101.

RNA extraction from WCDS and WTH was done on the 12 randomly selected cases. RIN means for those 12 WCDS were 6.3 +/-2.0, while WTH had a RIN mean of 6.5 +/-2.0. qRT-PCR results suggest that neuronal NEU-N and astrocyte GFAP RNA expression were not different between WCDS and WTH, while RNA expression of the well-known microglia protein IBA1 was upregulated in WCDS (Figure 3a). The same twelve cases were also used to do whole transcriptome sequencing and differential analyses, comparing Sorted Cells versus Homogenates. We classified the genes according to their specific cell expression signatures using a single cell mRNA database from the mouse cortex. Using these data, we defined an enrichment score based on deconvolution of the known relative expression of genes in different cell types, thus assigning each gene transcript to a specific cell (neuron, astrocyte, microglia, endothelial cell, and oligodendrocytes) or a “mixed” category when the expression was not specific to any cell type (Piras et al., 2020). We successfully sequenced more than 11,000 gene transcripts per WCDS. The average total mapped reads were 18,283,887 in the WCDS and 7,418,515 in WTH. Transcripts in WCDS and WTH included many that are specific for neurons (*n*= 626), astrocytes (*n*= 375), oligodendrocytes (*n*= 316), microglia (*n*= 684), endothelial cells (*n*= 578) and non-cell-specific transcripts (*n*=8,173; Fig 3b). When cell-specific WCDS transcripts expression were compared (*n*= 9,820) to WTH, several transcripts were upregulated or downregulated, but most numbers of cell-specific transcripts had a similar expression in both groups. The most evident differences were in microglia-specific genes, where almost 50% of the sequenced genes showed upregulation, and in neurons, where 40% of the neuron-specific genes were downregulated or expressed at lower levels in the dissociated cell preparations as compared to the whole cortical homogenates (Fig.4). Upregulation and downregulation were defined as 1.5 log2 fold change in either direction. Only a handful of genes reached a 5 log2 fold change and were microglia-specific.

**Figure 3.**
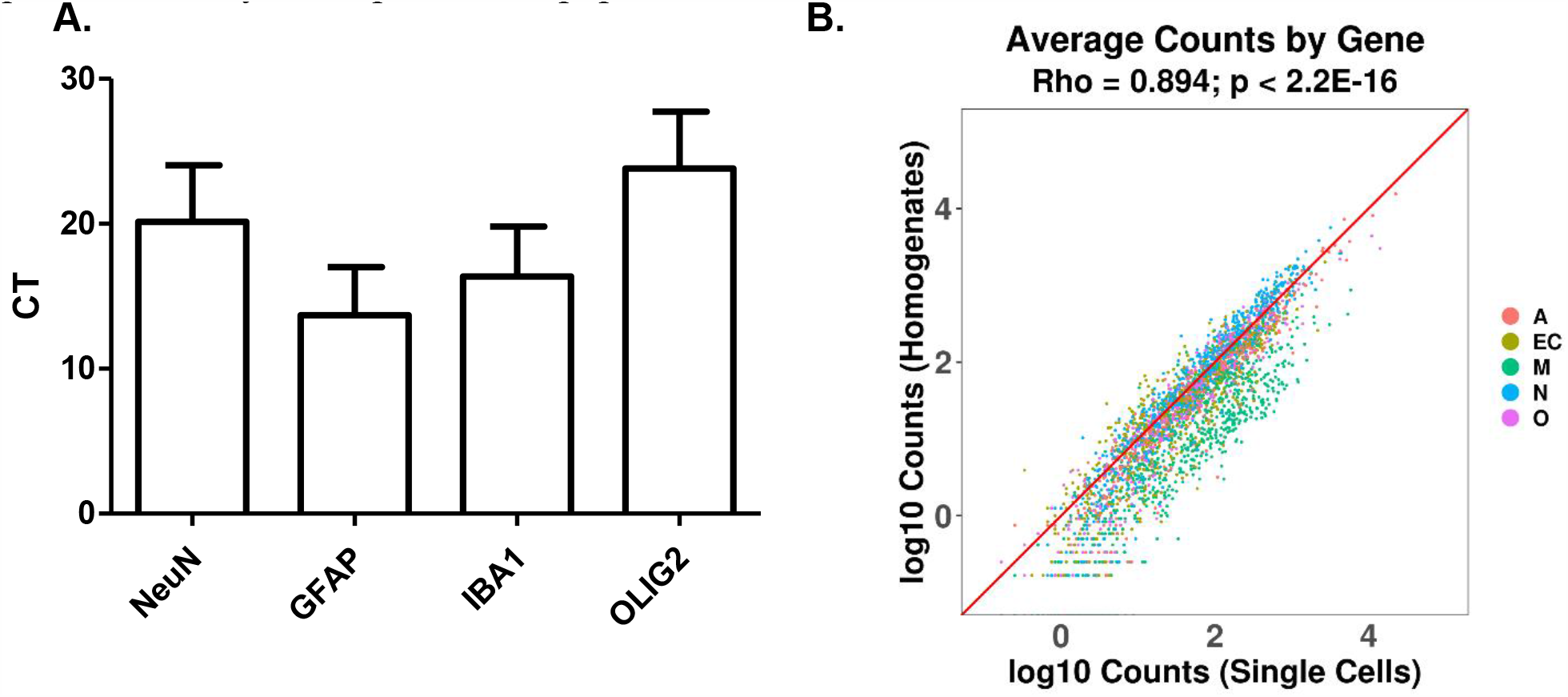
RNA transcript analysis of cell suspensions and adjacent intact brain tissue by qRT-PCR and RNAseq. qRT-PCR demonstrating the presence of transcripts for NeuN, GFAP, OLIG2, and Iba1 (A). WCDS express more than 11,000 different gene transcripts, including transcript specific for neurons (N), astrocytes (A), oligodendrocytes (O), microglia (M), and endothelial cells (E). When WCDS is compared to adjacent frozen whole tissue homogenates of the same cases, the total count of transcript per gene was similar, suggesting minimal transcript loss caused by the dissociation process (B).

**Figure 4.**
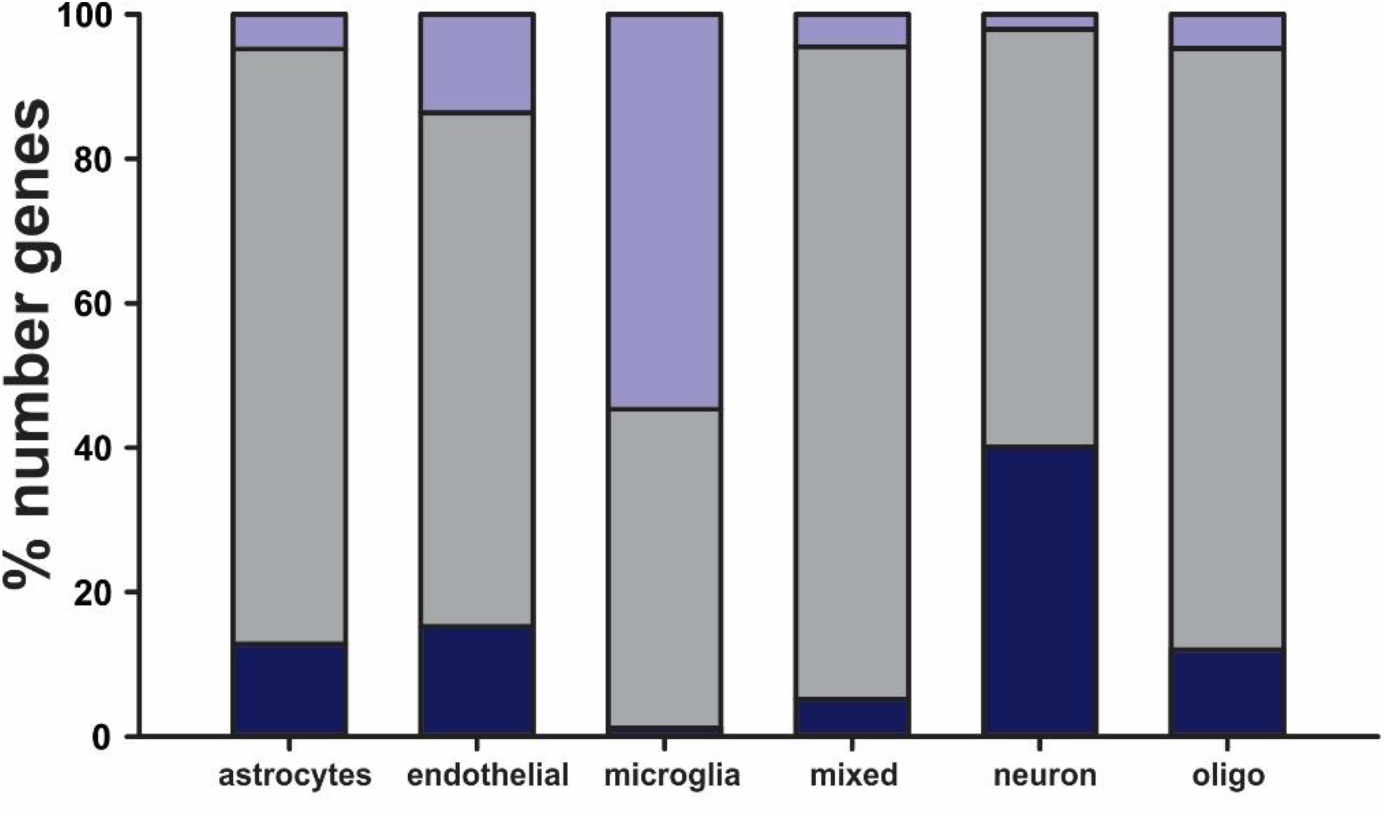
Sequenced transcripts from WCDS and WTH classified by cell expression signatures. Specific cell expression signatures using a single cell mRNA database from mouse cortex allow us to subdivide transcripts by cell-type or mixed (present in two or more cell types). For most cell types, transcript abundance was similar between WCDS and WTH (grey bar). Microglial-enriched suspension transcripts showed upregulation (purple) in almost 50% of the transcripts, while approximately 40% of neuron-specific transcripts were down-regulated (blue).

## Discussion

The scientific community now has many powerful tools to study genetic and transcriptomic changes in the human brain and disease. However, most studies using such techniques have been in whole-tissue homogenates; we now recognize that studying millions of various types of cells together, most with different transcriptomic profiles, could confound and obscure our understanding of the relevant functional pathways and regulations. LCM and single-nuclei methods have highlighted the importance of extending transcriptomic studies to a single-cell level by isolating phenotypically-defined populations to capture small changes that might be masked when compared with bulk tissue homogenizations (Srinivasan et al., 2020). Such studies have revolutionized the field, and even though there are already intriguing results published from single-nucleus studies of human brain cells, there have been very few similar studies of human whole-cell preparations. Results from different methodologies could complement each other; for example, single-nucleus preparations might result in more or quicker sequencing data than using LCM technology and is more widely accessible through archived frozen tissue, but nuclear mRNA is less abundant and may not represent the entire cellular complement. Having WCDS preparations allows us to study transcripts present in both nuclei and cytoplasm, without compromising quantity, speed, and quality of data generation.

In this study, we explored a new methodology to create WCDS at cold temperatures from a rapid-autopsy brain collection program that will serve as a new shared resource for researchers interested in cell population changes in aging and aging-related disorders. Transcriptional processes remain active at 37°C, therefore it has been hypothesized that cell dissociation in cold temperatures could limit gene expression artifact created by the tissue preparation methods, which might be expected to upregulate stress response genes (Fujita, 1999; O’Flanagan et al., 2019; Sonna, Fujita, Gaffin, & Lilly, 2002; Volovitz et al., 2016). To investigate this, we compared the transcriptome of twelve WCDS to WTH from the same cases to identify possible changes created by processing.

Our data shows that our suspensions contain relatively intact brain cells of several different types, with good yields of RNA and relatively good RIN values. We demonstrated that the RNA isolated from such suspensions is suitable for sequencing, and our results suggest that rather than losing transcripts during the WCDS processing, we captured a higher number of mapped transcripts than with the WTH samples. As this was surprising, we hypothesize that WCDS-RNA is derived mainly from cellular perikarya, which have more abundant transcripts than cell processes or neurites and would be a more substantial contributor to WTH-RNA. In this study we did not compare results from isolated nuclei, but we proposed that sequencing WCDS should contain higher transcript numbers per cell. Our results suggest that this new resource will be valuable and probably complementary to other resources to study cell-type-specific changes in aging and aging-related disorders. Some of these transcripts, when present in both WCDS and WTH preparations, were expressed at a higher or lower abundance in one source or the other. Overall, most transcripts were very similar between WCDS and WTH, but it is an important finding that almost 50% of the microglia-specific genes showed upregulation in the dissociated cell preparations when compared to the whole cortical homogenates. Even though our procedure is performed in a cold environment to reduce transcriptomic changes, microglia might still be reactive to the dissociation procedure, resulting in the upregulation of specific transcripts. Therefore, caution should be taken in interpreting future studies using isolated microglia.

## Data Availability

Data will be available by request and registration to www.brainandbodydonationprogram.org

## Acknowledgments

The Brain and Body Donation Program is supported by the National Institute on Aging (P30 AG19610 Arizona Alzheimer’s Disease Core Center), the Arizona Department of Health Services (contract 211002, Arizona Alzheimer’s Research Center), the Arizona Biomedical Research Commission (contracts 4001, 0011, 05-901 and 1001 to the Arizona Parkinson’s Disease Consortium) and the Michael J. Fox Foundation for Parkinson’s Research.

## Compliance with ethical standards

### Ethical approval

All subjects signed Institutional Review Board-approved informed consent, allowing both clinical assessments during life and several options for brain and organ donation after death.

